# The Relationship Between Household Cash Transfer Access and Cumulative Incidence of Diabetes and Prediabetes in Rural South Africa

**DOI:** 10.64898/2026.08.21.26361078

**Authors:** Maria Klein, Indrakshi Roy, Thomas Gaziano, Daniel Ohene-Kwofie, Evan Jordan, Corey Kalbaugh, Stephen Tollman, Molly Rosenberg

## Abstract

**Introduction:** Cash transfer programs could reduce diabetes risk by decreasing chronic stress and increasing food security, physical activity, and preventive care, but there is an evidence gap on the relationship between cash transfer access and diabetes and prediabetes incidence.

**Methods:** We used data from the “Health and Ageing in Africa: Longitudinal Studies in South Africa” (HAALSA) Indepth cohort of Black South Africans ages 40+ (N=5059). We fit log binomial models to estimate the relationship between household cash transfer eligibility (HCT) and cumulative incidence of diabetes and prediabetes between 2014/15 and 2021/22. We performed quantile regression to estimate change in continuous glucose values across the glucose distribution with additional HCT.

**Results:** No association was observed between HCT and diabetes risk. Each additional unit of HCT was associated with reduced prediabetes risk [aCIR (95% CI): 0.94 (0.90, 0.98); p=0.007]. The largest reduction in glucose values associated with additional HCT was at the highest end of the glucose distribution.

**Discussion:** Our observations suggest household cash transfer access reduces risk of prediabetes but not diabetes. However, household cash transfer access was associated with the largest decrease in glucose for the most severely glucose-impaired, implying the potential for HCT to reduce risk of hyperglycemic complications.

## INTRODUCTION

Diabetes is a condition in which the body cannot produce the hormone insulin (type 1) or cannot effectively use the insulin it produces (type 2), resulting in chronically high levels of blood sugar, or hyperglycemia.^1^ Diabetes can damage renal, nervous, and cardiovascular systems.^2^ The global burden of diabetes is burgeoning in terms of both prevalence and cost. By 2050, diabetes prevalence is estimated to grow from around 500 million to over 1.3 billion people^3^, and nearly $80 trillion (international dollars) will be spent on diabetes care between 2020 and 2050.^4^ Prediabetes, indicated by glucose levels above normal but below diabetes thresholds, affects conservatively another 464 million people and is also rapidly increasing.^5^ People with prediabetes may experience similar health complications and have high risk of future diabetes, with 5-10% converting to diabetes annually and up to a 70% lifetime conversion rate.^6^

The exact cause of diabetes is not yet fully understood, with evidence suggesting contributions from both modifiable and genetic components.^7^ However, 96% of diabetes cases are the potentially preventable and/or reversible type 2 form.^3^ Interventions addressing lifestyle factors have been widely successful at reducing the incidence of diabetes and reverting prediabetes to normoglycemia.^8–12^ Evidence-based lifestyle interventions for diabetes prevention include improving diet and nutrition, increasing physical activity, managing stress, and utilizing preventive care. Notably, lifestyle interventions are generally more effective than preventive medications such as metformin.^12^

Lower household income presents obstacles for all the aforementioned potential lifestyle levers.^13–20^ These obstacles manifest in earlier onset of diabetes and higher diabetes incidence in lower-income households.^17,21–23^ As such, increasing household income could address diabetes prevention challenges related to limited material resources.^24^

Household income can be intervened upon through cash transfer programs.^25^ Broadly defined, cash transfer programs are a type of social assistance program in which regular payments are made directly to recipients, often targeting vulnerable populations such as children or the elderly.^25,26^ Cash transfer programs can be conditional (i.e., requiring recipients to be compliant with behavioral conditions) or unconditional.^27^ The income provided by cash transfers could theoretically decrease chronic stress from poverty and increase food security, dietary diversity, recreational physical activity, and preventive care uptake, all of which are associated with reduced diabetes risk.^17,25,28^

The theoretical premise that cash transfer programs could reduce diabetes risk is supported by evidence in other health outcomes. Cash transfer programs are already documented to improve child and maternal health outcomes through increased food security and health services utilization.^25^ The same pathways could improve risk of diabetes, but cash transfer programs’ impact specific to diabetes is not well-studied.^17,25^ This gap is particularly apparent in low- and middle-income countries (LMICs), where cash transfer programs are often a central component of the social protection schema.^29^

With diabetes prevalence increasing globally, and growing fastest among LMICs^3^, there is an urgent need to understand if cash transfer programs can reduce the risk of diabetes in the general population and in LMICs specifically. To address this need, in this study we determined the association between cash transfer access and cumulative incidence of diabetes and prediabetes in a South African population.

## METHODS

### Parent Study and Setting

The HAALSA Indepth cohort is part of the Health and Ageing in Africa: Longitudinal Studies in South Africa portfolio of research jointly led by the University of Witwatersrand and the Harvard T.H. Chan School of Public Health. Eligibility criteria for HAALSA Indepth were: 1) age 40 years and older as of 1 July 2014, and 2) residing in the Agincourt Health and socio-Demographic Surveillance System (HDSS) study area for 12 months before the 2013 census update.^30^ Of 12,875 eligible adults in 2014, 6281 were randomly sampled using gender-specific sampling fractions to ensure a gender-balanced cohort.^30^ The final baseline sample included 5059 adults in 4393 households (86% response rate).^30^ Retention in HAALSA Indepth was high, with ∼94% of surviving respondents completing Wave 2 interviews in 2017/18 and Wave 3 interviews in 2021/22.

The Agincourt HDSS surveils a ∼450 km^2^ region of the largely rural Mpumalanga province in northeastern South Africa via an annual census of ∼120,000 people in approximately 22,000 households across 31 villages.^31^ The census includes an update of status and vital events involving every resident of the Agincourt sub-district. Fieldworkers visit each household in the study area to verify existing data including village and dwelling location, names, genders, ages, dates of births and death, identification of household heads and familial structures, details of in-migration and out-migration, and marriage and union status, as well as record any new event experienced by each household member.

The Agincourt sub-district itself is a former ‘homeland’ region, where xiTsonga-speaking Black South Africans were forcibly moved as part of legislated racial segregation during Apartheid from 1948-1993.^31^ During the Apartheid era, the Black population in the region had low access to education, poor public services, and high unemployment. Historically, available jobs have been low-paying and in mining, farming, and domestic work.^32,33^ Economic development in the Agincourt sub-district has improved since the end of Apartheid, but unemployment remains high and gaps persist in health services and food security.^31^

In terms of health services, approximately 80% of the South African population accesses health care through government-sponsored clinics where primary care services are delivered free of charge to any resident.^34–36^ In the Agincourt sub-district, the primary health care system consists of six clinics, two health centers, and three district hospitals within 60 km.^31^ In terms of food security, the diet of Agincourt-area residents and of South Africans more broadly are typically low in fruit and vegetable consumption^37^ and, particularly among cash transfer recipient households, are high in starchy carbohydrates.^38^ Among South African cash transfer program participants such as those in the Agincourt sub-district, benefits are often pooled at the household level and spent predominantly on food.^39^ Even so, many recipients state that their diets are still nutritionally inadequate, energy-dense, and lacking in diversity.^38^

### Ethical Considerations

The HAALSA Indepth study received ethical approvals from Mpumalanga Provincial Research and Ethics Committee, University of the Witwatersrand Human Research Ethics Committee (ref. M141159), and the Harvard T.H. Chan School of Public Health, Office of Human Research Administration (ref. C13–1608–02).^30^ The Indiana University Institutional Review Board deemed this analysis not human subjects research (ref. 28496).

### Analytic Sample

From the full HAALSA Indepth cohort (N=5059), we excluded 65 participants (1.3%) for whom we were unable to determine HCT eligibility. We could not determine HCT eligibility for these individuals due to meaningful discrepancies in age reported in HAALSA Indepth and age reported in the AHDSS census. As our exposure construction relied on age to determine eligibility for cash transfer benefits, we excluded these participants rather than risk misclassification of their true benefit eligibility. We also excluded 414 participants (8.1%) for whom we could not establish glucose impairment status due to missing Wave 1 blood glucose values. Our resulting study sample consisted of 4580 participants, or 90.5% of the full HAALSA Indepth cohort (**Figure 1**).

**Figure 1.**
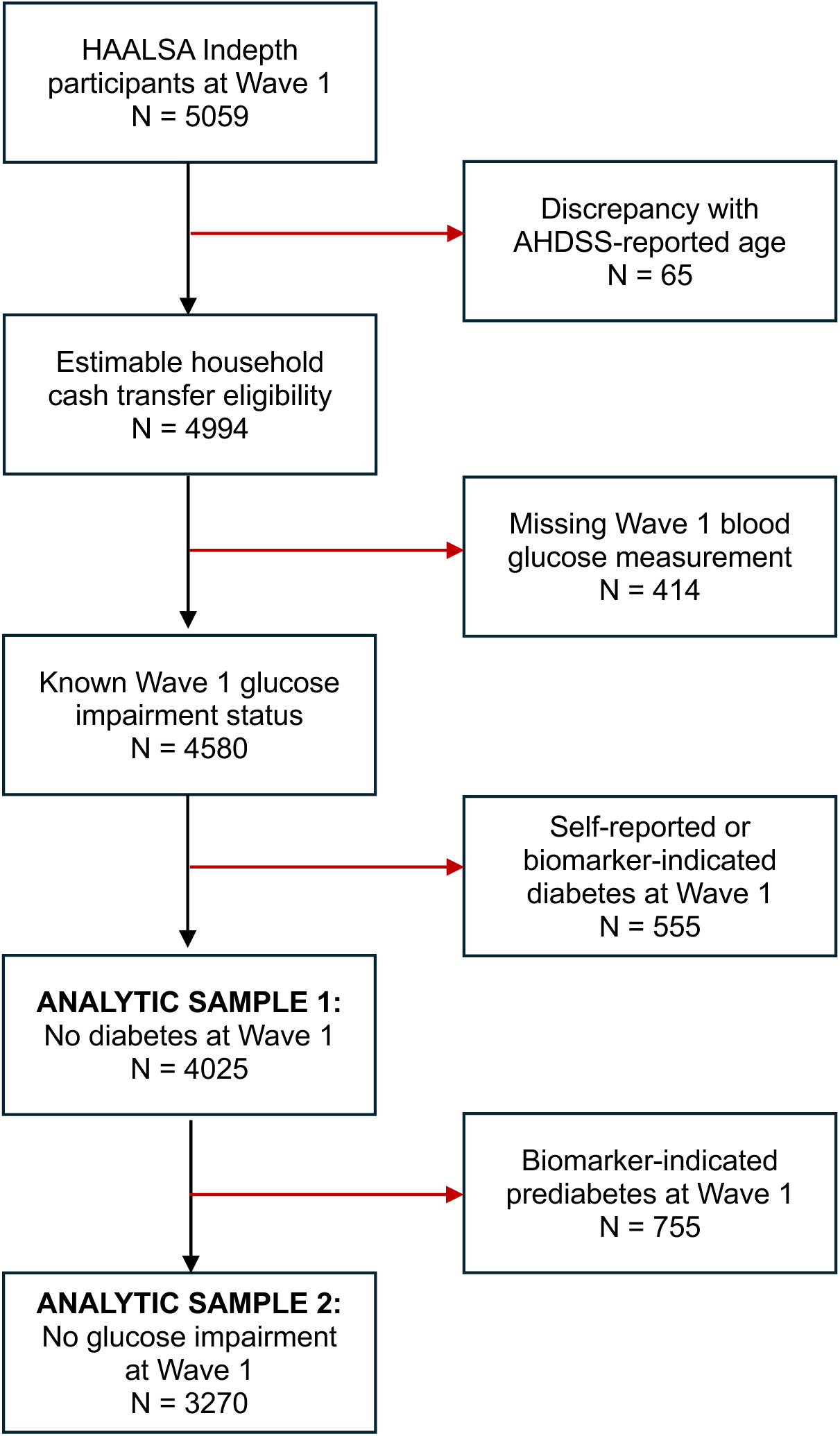
Analytic sample construction.

From our study sample, we derived 2 primary analytic samples: participants without self-reported or biomarker-indicated diabetes at Wave 1 (N=4025) and participants without self-reported diabetes or biomarker-indicated glucose impairment at Wave 1 (N=3270). Self-reported diabetes was defined as participants who reported they had ever been told by a doctor, nurse, or other healthcare worker that they had raised blood sugar or diabetes (exclusive of pregnancy for women) at their HAALSA Indepth Wave 1 interview. Biomarker-indicated diabetes was defined as a point-of-care blood glucose measurement at or above diabetes diagnostic thresholds (≥ 7.0 mmol/L when fasted 8 hours or more, ≥ 11.1 mmol/L when nonfasted) collected at Wave 1 interview.^40,41^ Biomarker-indicated glucose impairment was defined as a point-of-care blood glucose measurement of ≥ 5.6 mmol/L fasted or ≥ 7.8 mmol/L random collected at Wave 1 interview.^40^ Biomarker-indicated glucose impairment includes participants with glucose measurements falling in the prediabetes range in addition to those at or above diabetes thresholds.

Although diabetes and prediabetes thresholds were developed for plasma glucose and HAALSA Indepth measured blood glucose levels, the CareSens N point-of-care devices used by HAALSA Indepth were calibrated to provide plasma-equivalent readings^42^, and these thresholds have been previously used to assign diabetes status in the HAALSA Indepth population.^43^

### Key Measures

#### Exposure: household cash transfer

Our exposure was the average monthly household cash transfer (HCT) available to HAALSA Indepth participants between 2000 and their Wave 1 interview (2014/2015). To create this variable, we used AHDSS census rosters to construct cash transfer eligibility histories for each member of a HAALSA Indepth participant’s household by month for the two largest South African cash transfer programs: the Older Person’s Grant (OPG) and the Child Support Grant (CSG).

The OPG and the CSG are means-tested but otherwise unconditional cash transfer programs designed to alleviate poverty among vulnerable South African populations. The OPG was introduced in 1928 but Black South Africans have only been eligible for full benefits since 1993.^44^ Historically, women were OPG-eligible at age 60, while men were not eligible until 65. Eligibility expanded to age 60 for men between 2008 and 2010.^45^ As of April 2026, the monthly OPG grant was R2400 for ages 60 to 74 (∼$142 USD) and R2420 for ages 75 and older.^46^ The CSG was established in 1998 to help offset the costs of raising children, typically paid to the child’s biological mother.^47,48^ The grant was originally offered to households with children up to age 7, but expanded between 2003 and 2012 to cover children up to age 18.^48^ As of April 2026, the monthly CSG grant was R580 (∼$34 USD) per age-eligible child.^46^

We populated household members’ OPG and CSG eligibility periods with the South African Social Security Agency (SASSA)-verified historical monthly benefit values in South African Rand, then aggregated individual values by household for each month, adjusting for inflation using the 2023 South African Consumer Price Index (base index = December 2021).^49^ To account for differences in household sizes and changing household structures over time, each month’s household total was divided by the number of individuals living in the household that month to yield a per capita monthly household cash transfer eligibility amount. Each month’s per capita value was summed, resulting in a cumulative household cash transfer amount (per capita) over the entire exposure accrual period. Finally, each cumulative total was divided by the number of months the household was present in the AHDSS surveillance area, resulting in the average monthly household cash transfer amount (per capita), heretofore called “HCT.”

Although AHDSS household rosters extend back to March 1992, we chose to begin our exposure accumulation in January 2000 for two reasons. First, sources of potential variation in HCT amount between 1993 and 1998 are more limited, as the CSG program did not begin until 1998 and the first expansions to program eligibility for CSG or OPG did not begin until 2003. Second, uptake to CSG was initially low^50^, meaning eligibility for benefits was less likely to approximate receipt of benefits in the CSG program’s earliest stages.

#### Outcomes: cumulative incidences of diabetes, prediabetes, and any glucose impairment

Our primary outcomes were the cumulative incidences of 1) diabetes alone, 2) prediabetes alone, and 3) any glucose impairment (diabetes and prediabetes combined) between Waves 1 (2014/15) and 3 (2021/22). Using the analytic sample of HAALSA Indepth participants without diabetes at Wave 1, cumulative incidence of diabetes was defined as self-reported diabetes and/or point-of-care blood glucose measurement of ≥ 7.0 mmol/L fasted or ≥ 11.1 mmol/L random at Wave 3.^40,41^ Using the analytic sample without glucose impairment at Wave 1, cumulative incidence of prediabetes was defined as a point-of-care blood glucose measurement of ≥ 5.6 mmol/L fasted and 7.0 mmol/L fasted or ≥ 7.8 mmol/L and < 11.1 mmol/L random^40^ but no self-reported diabetes at Wave 3. Using the analytic sample without any glucose impairment at Wave 1, cumulative incidence of any glucose impairment was defined as a point-of-care blood glucose measurement of ≥ 5.6 mmol/L fasted or ≥ 7.8 mmol/L random^40^ and/or self-reported diabetes at Wave 3.

#### Covariates

Additional variables were used to adjust for confounding, contextualize our study population, and inform a multiple imputation sensitivity analysis. The socioeconomic profile of the Agincourt area suggests that all HAALSA Indepth households meet the means-testing requirements for the OPG and CSG programs.^30^ As such, age and sex were the only two structural predictors of individual OPG and CSG eligibility, the component programs we used to operationalize HCT. At the household level, number of household members was also a structural predictor of amount of household cash transfer eligibility. To address this structural predictor, we built a per capita adjustment into our HCT construction. This per capita adjustment allowed us to account for differences in household size without restricting some of the potential sources of exposure variation, such as number of children or number of pension-eligible adults. With this household size adjustment, we established age (in continuous years) and sex (man or woman) as a minimally sufficient adjustment set for modeling the relationships between HCT and cumulative incidence of diabetes and prediabetes. For exploratory stratifications by age, we also categorized age into groups of 40 to 59 years, 60 to 79 years, and 80 years and older.

To contextualize our study population, we have reported household wealth quintile (1 lowest, 5 highest), average number of children, education level (no formal education or any formal education), average number of children, country of birth (South Africa or other), and marital status (currently partnered or not currently partnered). Household wealth was assessed by the self-reported or observed presence or absence of 26 household assets (e.g., housing characteristics, vehicles, and livestock)^51,52^; asset scores were converted into wealth index quintiles using principal components analysis.^30^

To address the ∼8% and ∼28% of missing point-of-care blood glucose measurements at Wave 1 and 3, respectively, we selected additional HAALSA Indepth covariates predictive of blood glucose values to inform a multiple imputation model. Predictive covariates included Wave 1 HbA1c, number of hours since last eaten at the time of Wave 1 and Wave 3 glucose measurements, Wave 1 and Wave 3 self-reported diabetes diagnoses (yes or no), self-reports of diabetes treatment at Wave 1 and Wave 3 (yes or no), and Wave 1 and Wave 3 waist-to-hip ratios.

### Statistical analysis

We estimated Wave 1 prevalence and cumulative incidence by Wave 3 with log-binomial models specified with cluster-robust standard errors (CRSE). We chose to use CRSE because it was possible for households to contain more than one HAALSA Indepth participant. However, 86% of our households contained only one participant, making within household variation difficult to estimate. CRSE allowed us to account for any residual correlation within households without the problems that a multilevel model may encounter given the near-zero variance in the household-level cluster.^53^ We used inverse probability weights in models estimating cumulative incidence between Waves 1 and 3 to account for mortality, attrition, and refusal of point-of-care measurement.

To estimate the relationship between continuous HCT and the cumulative incidences of diabetes, prediabetes, and any glucose impairment, we again used log-binomial models with CRSE. We substituted modified Poisson when log-binomial models failed to converge.^54^ Models were specified with inverse probability weights to mitigate possible bias due to differential rates of death, loss to follow up, and refusal of measurement. We fit unadjusted models and models adjusted for age (centered around each group’s mean) and sex. To increase interpretability, we scaled HCT in units of R100 and centered it around its mean. Beta coefficients extracted from the models were exponentiated to yield risk ratios, estimating the relative change in cumulative incidence associated with each additional R100 of HCT. We employed the same modeling strategy for exploratory analyses within sex and age group strata. We additionally specified a quantile regression model to estimate the change in blood glucose values associated with an additional R100 of HCT at the 10^th^, 25^th^, 50^th^, 75^th^, and 90^th^ percentile of Wave 3 glucose distribution.

We tested the robustness of our results through several sensitivity analyses. The first sensitivity analysis tested use of World Health Organization (WHO) criteria for establishing presence of prediabetes rather than the American Diabetes Association criteria used in our main analysis. WHO criteria are slightly more conservative, recommending a fasted threshold of 6.1 mmol/L instead of 5.6 mmol/L.^55^ We also tested alternative operationalizations of our HCT exposure. The first alternative operationalization removed our adjustment for the AHDSS census expansion. 11 villages have been added to the AHDSS surveillance area since 2007. Newly surveilled households would have missing household roster data (necessary for constructing HCT) between the start of our HCT accumulation period (year 2000) and the date the village was added to the census. We backfilled this missing data based on the household’s roster at the date their village was first surveilled, accounting for children’s birthdates so that they were only backfilled as household members until their date of birth. The second alternative operationalization started our exposure accumulation period at the AHDSS census start date of March 1992 instead of January 2000. Our third alternative operationalization used total cumulative HCT (per capita) instead of average monthly HCT. Total cumulative HCT removes the adjustment for length of residence in the AHDSS surveillance area. Our final sensitivity analysis addressed the ∼8% and ∼28% of glucose values missing at Waves 1 and 3, respectively. We used multiple imputation by chained equation (MICE) to impute missing glucose values. We defined the MICE predictor matrix such that number of hours last ate only informed the imputation of blood glucose values at the same wave and individual and household identifiers were excluded as predictors for any variable. We set the MICE methods to use two-level predictive mean matching (2l.pmm) for any variable with a non-zero intraclass correlation coefficient; variables with an intraclass correlation coefficient of ∼zero used predictive mean matching (pmm).

Significance levels for all test statistics were set at alpha = 0.05 unless otherwise noted. Statistical analyses were performed in RStudio version 2026.01.1+403.^56^ Log-binomial and modified Poisson models were fit using the base R ‘stats’ package (version 4.5.3) and quantile regression models were fit using the ‘quantreg’ package (version 6.1).^57^ The ‘mice’ package (version 3.19.0) was used for the multiple imputation sensitivity analysis.^58^

## RESULTS

### Sample characteristics

The full study sample had a mean age of 62.4 years and a mean HCT of R368.8 (∼$21 USD) (**Table 1**). Between 2014/15 and 2021/22, there were 669 cases of incident glucose impairment, 386 cases of incident prediabetes, and 374 cases of incident diabetes. On average, participants with incident glucose impairment, incident prediabetes alone, and incident diabetes alone lived in households with less HCT (R320.3, R321.7, and R313.6, respectively) than all study sample participants. Participants with incident glucose impairment tended to be younger and have more education than participants who did not develop glucose impairment. Participants with incident prediabetes alone also tended to be younger and have more education than participants who did not develop prediabetes. Participants with incident diabetes alone tended to have more children than participants who did not develop diabetes. Comparison data for outcome groups are available in supplemental tables S1 and S2.

**Table 1.** Study sample characteristics: HAALSA Indepth participants, 2014/15.

| Sample Characteristic<sup>1</sup> | Full study sample<br>N = 4580 | Incident glucose impairment<sup>2</sup><br>N = 669 | Incident prediabetes alone<sup>3</sup><br>N = 386 | Incident diabetes alone<sup>4</sup><br>N = 374 |
| --- | --- | --- | --- | --- |
| HCT in Rand<sup>5</sup> | 368.8 (347.5) | 320.3 (283.0) | 321.7 (285.4) | 313.6 (265.2) |
| Age in years [min, max] | 62.4 [40, 112] | 61.2 (40, 95] | 61.5 [40, 95] | 60.9 [40, 94] |
| Age group |  |  |  |  |
| 40 to 59 years | 2022 (44%) | 313 (47%) | 182 (47%) | 175 (47%) |
| 60 to 79 years | 2042 (45%) | 316 (47%) | 183 (47%) | 175 (47%) |
| 80+ years | 516 (11%) | 40 (6%) | 21 (5%) | 24 (6%) |
| Women | 2467 (54%) | 390 (58%) | 221 (57%) | 230 (62%) |
| Number of children | 4.6 (2.8) | 4.8 (2.8) | 4.7 (2.7) | 4.9 (2.8) |
| Household wealth index quintile<sup>6</sup> |  |  |  |  |
| 1 | 946 (21%) | 138 (21%) | 80 (21%) | 72 (19%) |
| 2 | 909 (20%) | 132 (20%) | 74 (19%) | 66 (18%) |
| 3 | 897 (20%) | 137 (21%) | 78 (20%) | 75 (20%) |
| 4 | 904 (20%) | 133 (20%) | 74 (19%) | 84 (23%) |
| 5 | 924 (20%) | 129 (19%) | 80 (21%) | 77 (21%) |
| No formal education | 2087 (46%) | 308 (46%) | 183 (47%) | 159 (43%) |
| missing | 12 | 1 | 0 | 1 |
| Born in South Africa | 3186 (70%) | 448 (67%) | 250 (65%) | 265 (71%) |
| missing | 4 | 0 | 0 | 0 |
| Currently partnered | 2351 (51%) | 377 (56%) | 224 (58%) | 205 (55%) |
| missing | 2 | 0 | 0 | 0 |

<sup>1</sup>Mean (SD), N (%) unless otherwise noted. Percentages calculated from non-missing totals; may not total 100% due to rounding.
<sup>2</sup>In normoglycemic sample at Wave 1, self-reported diabetes or point-of-care glucose measurement $\geq 5.6$ mmol/L fasted or $\geq 7.8$ mmol/L random at Wave 3.
<sup>3</sup>In normoglycemic sample at Wave 1, point-of-care glucose measurement $\geq 5.6$ mmol/L but $< 7.0$ mmol/L fasted or $\geq 7.8$ mmol/L but $< 11.1$ mmol/L random.
<sup>4</sup>In sample without diabetes at Wave 1, self-reported diabetes and point-of-care glucose measurement $\geq 7.0$ mmol/L fasted or $11.1$ mmol/L random.
<sup>5</sup>HCT = household cash transfer. The average cash transfer amount available to HAALSA Indepth participant's household (per capita) each month between 2000 and Wave 1 interview.
<sup>6</sup>Household wealth index based on household asset index score. Median raw asset score for full HAALSA Indepth cohort was -0.453.

### Prevalence and cumulative incidence of diabetes, prediabetes, and any glucose impairment

In 2014/15, diabetes was present in 12.1% and prediabetes in 16.5% of the full study sample, resulting in a prevalence of any impaired glucose of 28.6% (**Table 2**). When comparing within age and sex strata, Wave 1 prevalence was lower in men than women and among those aged 40 to 59 compared to older age groups (**Figure 2**).

**Table 2.** Prevalence of glucose impairment: HAALSA Indepth, 2014/15.

|  | Any glucose impairment<sup>1</sup> |  | Diabetes alone |  | Prediabetes alone |  |
| --- | --- | --- | --- | --- | --- | --- |
|  | Prev | 95% CI | Prev | 95% CI | Prev | 95% CI |
| Total population | 28.6% | 27.3, 29.9 | 12.1% | 11.2, 13.1 | 16.5% | 15.4, 17.6 |
| By sex |  |  |  |  |  |  |
| Men | 25.4% | 23.5, 27.3 | 11.2% | 9.9, 12.6 | 14.2% | 12.7, 15.7 |
| Women | 31.4% | 29.6, 33.2 | 12.9% | 11.6, 14.3 | 18.5% | 17.0, 20.0 |
| By age group |  |  |  |  |  |  |
| 40 to 59 years | 23.5% | 21.7, 25.4 | 9.1% | 7.9, 10.4 | 14.4% | 13.0, 16.0 |
| 60 to 79 years | 32.6% | 30.6, 34.7 | 14.8% | 13.3, 16.4 | 17.8% | 16.2, 19.5 |
| 80+ years | 32.8% | 28.8, 36.9 | 13.4% | 10.6, 16.5 | 19.4% | 16.1, 22.9 |

<sup>1</sup>Any glucose impairment = prediabetes and diabetes combined

**Figure 2.**
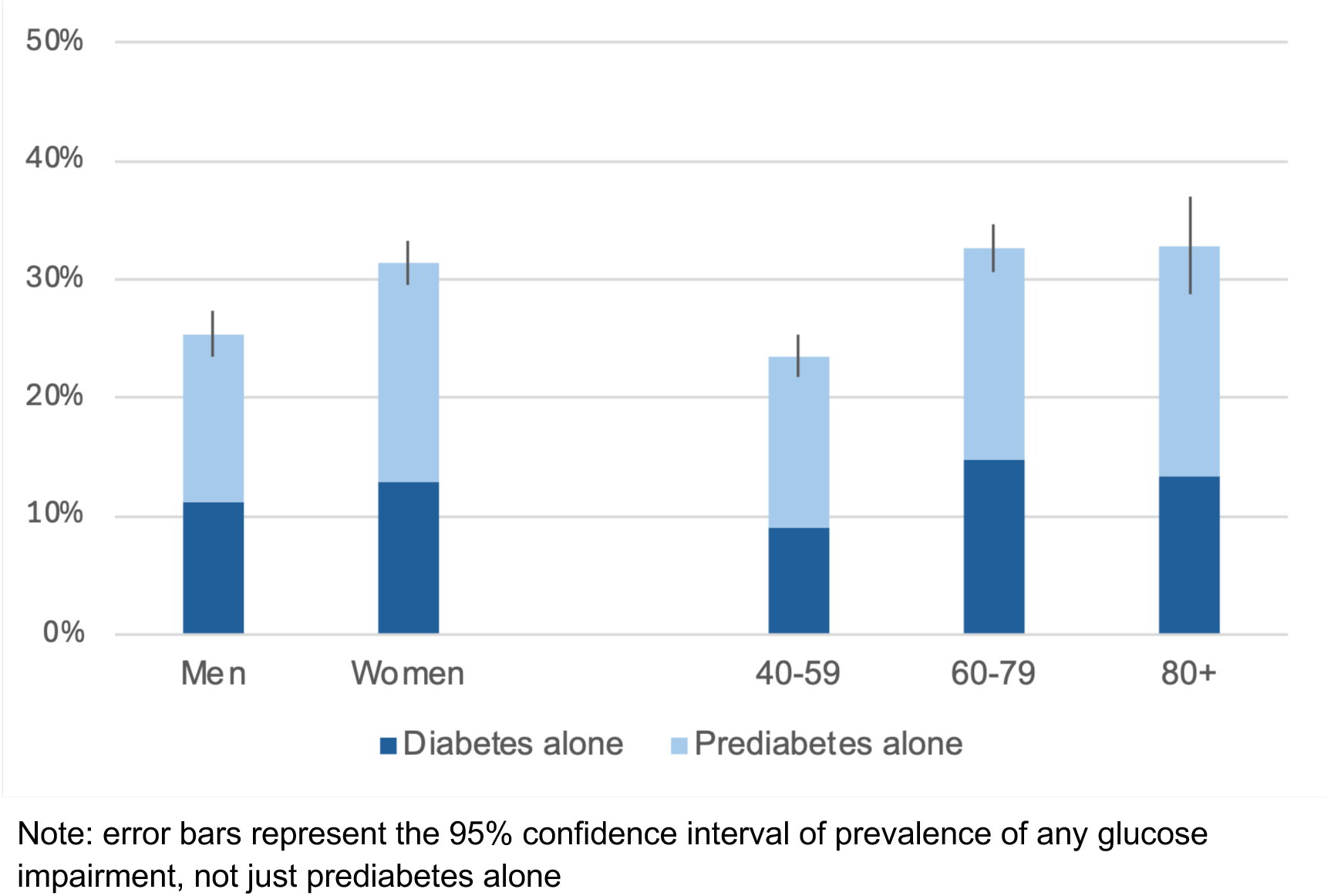
Prevalence of glucose impairment: HAALSA Indepth, 2014/15.

By 2021/22, cumulative incidence of diabetes and prediabetes was 16.8% and 21.7%, respectively (**Table 3**). The cumulative incidence of any glucose impairment was 37.5%. The risk set for incident diabetes was participants with no diabetes at Wave 1 and the risk set for incident prediabetes was restricted to normoglycemic individuals only. Accordingly, these two estimates are not directly additive, i.e., the sum of the cumulative incidences of prediabetes and diabetes does not equal the cumulative incidence of any glucose impairment. When comparing within age and sex strata, cumulative incidence was higher in women than men for any glucose impairment. Cumulative incidence was higher in ages 40 to 59 than in older ages for diabetes (although only statistically significant for ages 60 to 79), and lower in ages 40 to 59 than in older ages for diabetes (although again only statistically significant for ages 60 to 79).

**Table 3.**
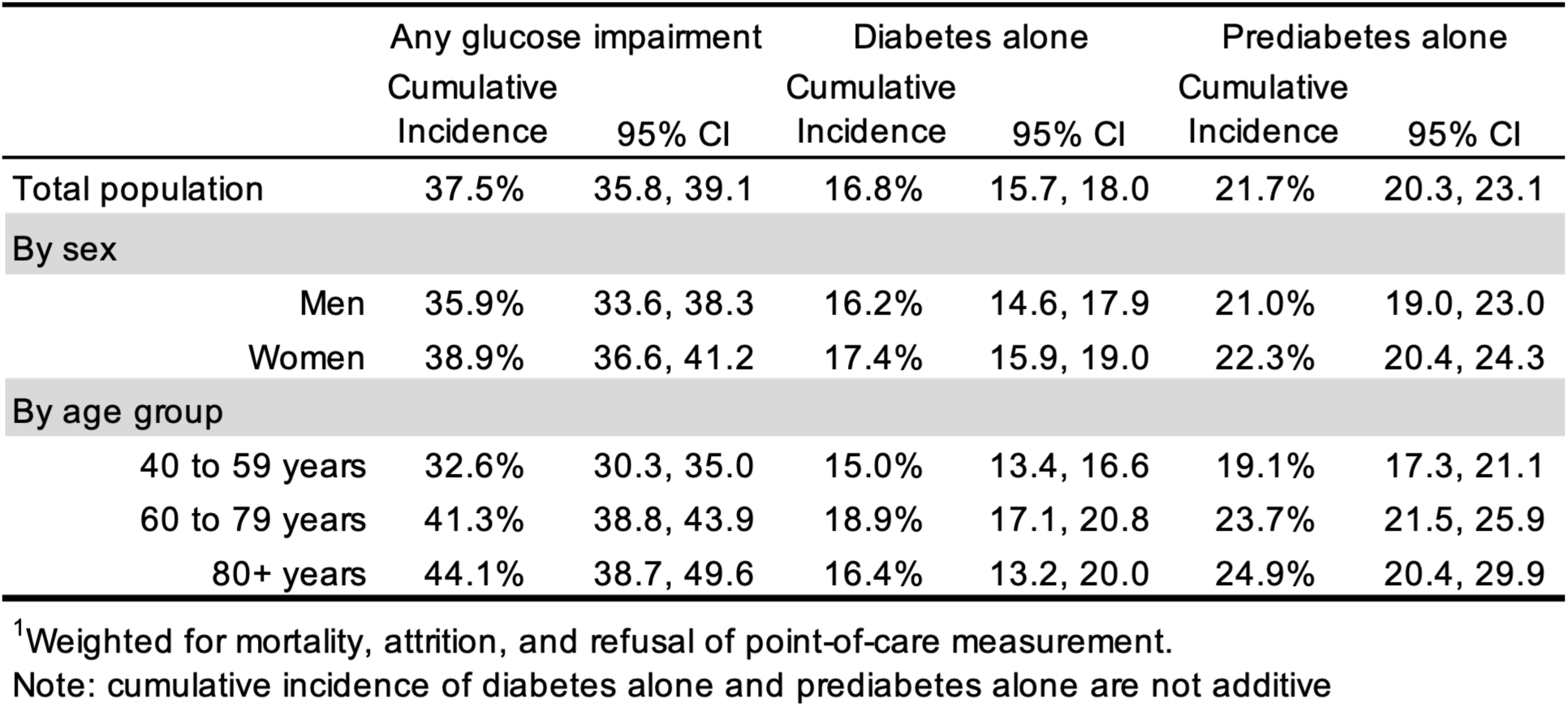
Cumulative incidence ^1^ of glucose impairment between 2014/15 and 2021/22: HAALSA Indepth.

|  | Any glucose impairment |  | Diabetes alone |  | Prediabetes alone |  |
| --- | --- | --- | --- | --- | --- | --- |
|  | Cumulative Incidence | 95% CI | Cumulative Incidence | 95% CI | Cumulative Incidence | 95% CI |
| Total population | 37.5% | 35.8, 39.1 | 16.8% | 15.7, 18.0 | 21.7% | 20.3, 23.1 |
| By sex |  |  |  |  |  |  |
| Men | 35.9% | 33.6, 38.3 | 16.2% | 14.6, 17.9 | 21.0% | 19.0, 23.0 |
| Women | 38.9% | 36.6, 41.2 | 17.4% | 15.9, 19.0 | 22.3% | 20.4, 24.3 |
| By age group |  |  |  |  |  |  |
| 40 to 59 years | 32.6% | 30.3, 35.0 | 15.0% | 13.4, 16.6 | 19.1% | 17.3, 21.1 |
| 60 to 79 years | 41.3% | 38.8, 43.9 | 18.9% | 17.1, 20.8 | 23.7% | 21.5, 25.9 |
| 80+ years | 44.1% | 38.7, 49.6 | 16.4% | 13.2, 20.0 | 24.9% | 20.4, 29.9 |

<sup>1</sup>Weighted for mortality, attrition, and refusal of point-of-care measurement.
Note: cumulative incidence of diabetes alone and prediabetes alone are not additive

### HCT and cumulative incidence of diabetes, prediabetes, and any glucose impairment

No association was observed between HCT and cumulative incidence of any glucose impairment in either unadjusted or models adjusted for age and sex (**Table 4**). Likewise, no associations were observed among HCT and cumulative incidence of diabetes. When adjusted for age and sex, HCT was associated with a reduced risk of prediabetes [aCIR (95% CI): 0.94 (0.90, 0.98); p=0.007]. For each additional R100 of HCT, risk of prediabetes by Wave 3 decreased 6%.

**Table 4.** Association between HCT and cumulative incidence of glucose impairment: HAALSA Indepth.

|  | Any glucose impairment |  |  | Diabetes alone |  |  | Prediabetes alone |  |  |
| --- | --- | --- | --- | --- | --- | --- | --- | --- | --- |
|  | CIR | 95% CI | P value | CIR | 95% CI | P value | CIR | 95% CI | P value |
| Full population |  |  |  |  |  |  |  |  |  |
| Unadjusted | 1.01 | 0.98, 1.03 | 0.659 | 1.01 | 0.96, 1.06 | 0.788 | 0.99 | 0.96, 1.02 | 0.484 |
| Adjusted<sup>1</sup> | 0.98 | 0.94, 1.02 | 0.282 | 1.00 | 0.93, 1.07 | 0.908 | **0.94** | **0.90, 0.98** | **0.007** |
| By sex<sup>2</sup> |  |  |  |  |  |  |  |  |  |
| Men | 0.99 | 0.93, 1.04 | 0.588 | 1.01 | 0.92, 1.10 | 0.814 | **0.94** | **0.89, 0.99** | **0.029** |
| Women | 0.96 | 0.92, 1.01 | 0.120 | 0.97 | 0.90, 1.04 | 0.384 | 0.95 | 0.88, 1.01 | 0.117 |
| By age<sup>1</sup> |  |  |  |  |  |  |  |  |  |
| 40 to 59 years | 0.95 | 0.89, 1.03 | 0.214 | 1.02 | 0.93, 1.12 | 0.740 | 0.90 | 0.80, 1.02 | 0.096 |
| 60 to 79 years | 0.99 | 0.94, 1.04 | 0.722 | 1.01 | 0.92, 1.11 | 0.844 | 0.96 | 0.91, 1.01 | 0.082 |
| 80+ years | 0.98 | 0.90, 1.06 | 0.594 | 0.99 | 0.87, 1.13 | 0.846 | 0.94 | 0.83, 1.07 | 0.369 |

<sup>1</sup>Adjusted for sex and continuous age.
<sup>2</sup>Adjusted for continuous age.

We observed little effect modification in exploratory models stratified by age and sex (**Table 4**). No associations were observed between HCT and cumulative incidence of any glucose impairment, with similarly sized point estimates consistently below the null across all strata. We also observed no associations within age and sex strata for cumulative incidence of diabetes. HCT was associated with a reduction of risk of prediabetes in men [aCIR (95% CI): 0.94 (0.89, 0.99); p=0.029]. Women had a comparably sized point estimate [aCIR (95% CI): 0.95 (0.88, 1.01); p=0.117], though not statistically significant. Across age groups, HCT trended toward the largest reduction of risk in prediabetes among ages 40 to 59 years [aCIR (95% CI): 0.90 (0.80, 1.02); p=0.096], though point estimates were only marginally significant for the 40 to 59 years and the 60 to 79 years age groups and were not statistically significant for ages 80 and older.

### Quantile regression

Quantile regression at the 10^th^, 25^th^, 50^th^, 75^th^ and 90^th^ percentiles show point estimates below the null for each quantile, suggesting that increased HCT is associated with a reduction in glucose values regardless of whether values were at the normoglycemic end or the impaired end of the Wave 3 glucose distribution (**Figure 3**). The reduction in glucose values associated with HCT is largest at the highest end of the glucose distribution. The decrease in glucose levels associated with additional HCT at the 75^th^ percentile [aβ (95% CI): -0.074 (-0.101, -0.014)] is nearly double the decrease associated with the conditional mean [aβ (95% CI): -0.043 (-0.082, - 0.004)]; at the 90^th^ percentile, the decrease is more than threefold [aβ (95% CI): -0.150 (-0.185, - 0.017)] (**Table 5**).

**Figure 3.**
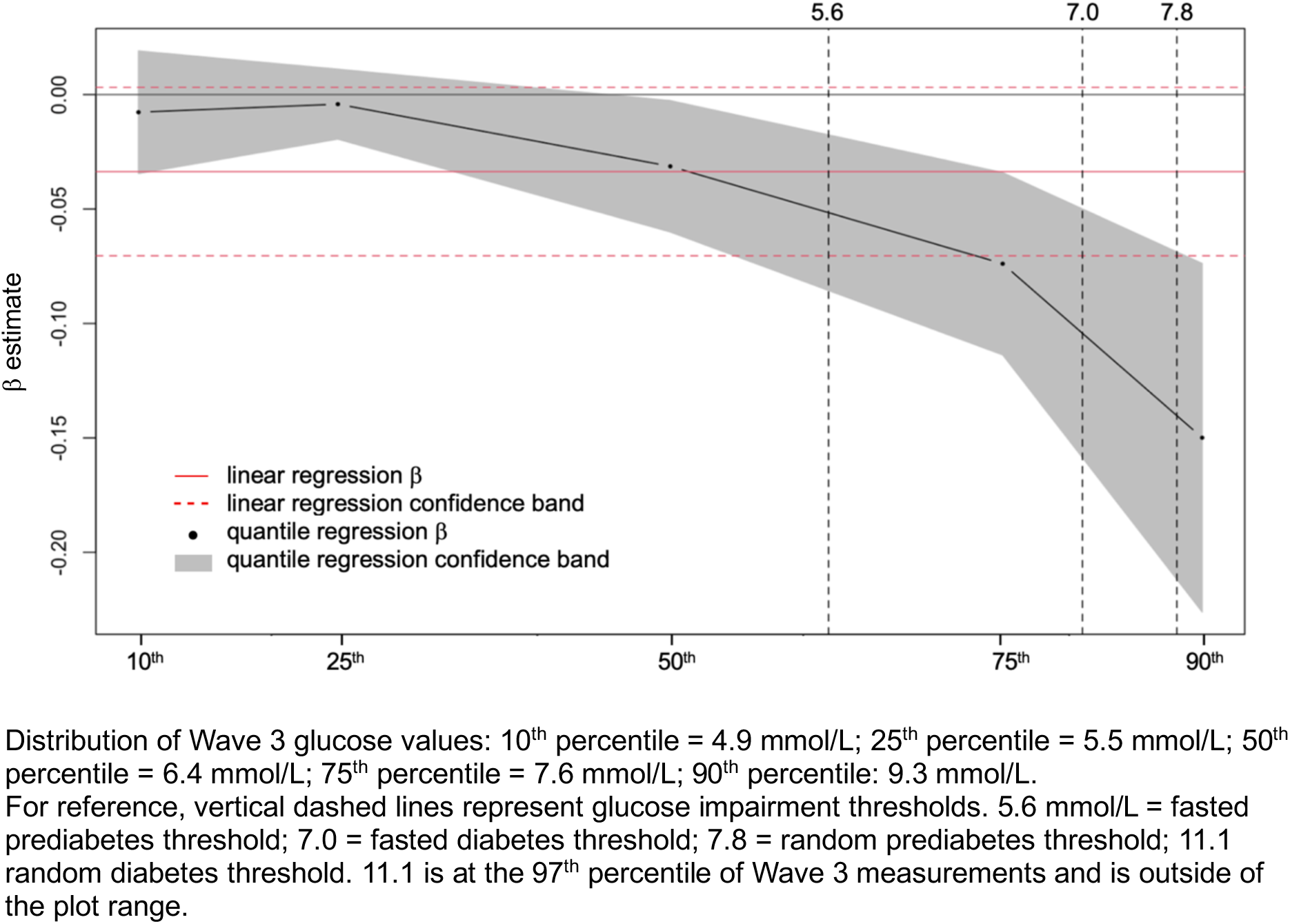
Change in Wave 3 glucose values associated with 1-unit increase of HCT in the Wave 1 normoglycemic HAALSA Indepth population, across quantiles of blood glucose measurements in mmol/L.

**Table 5.**
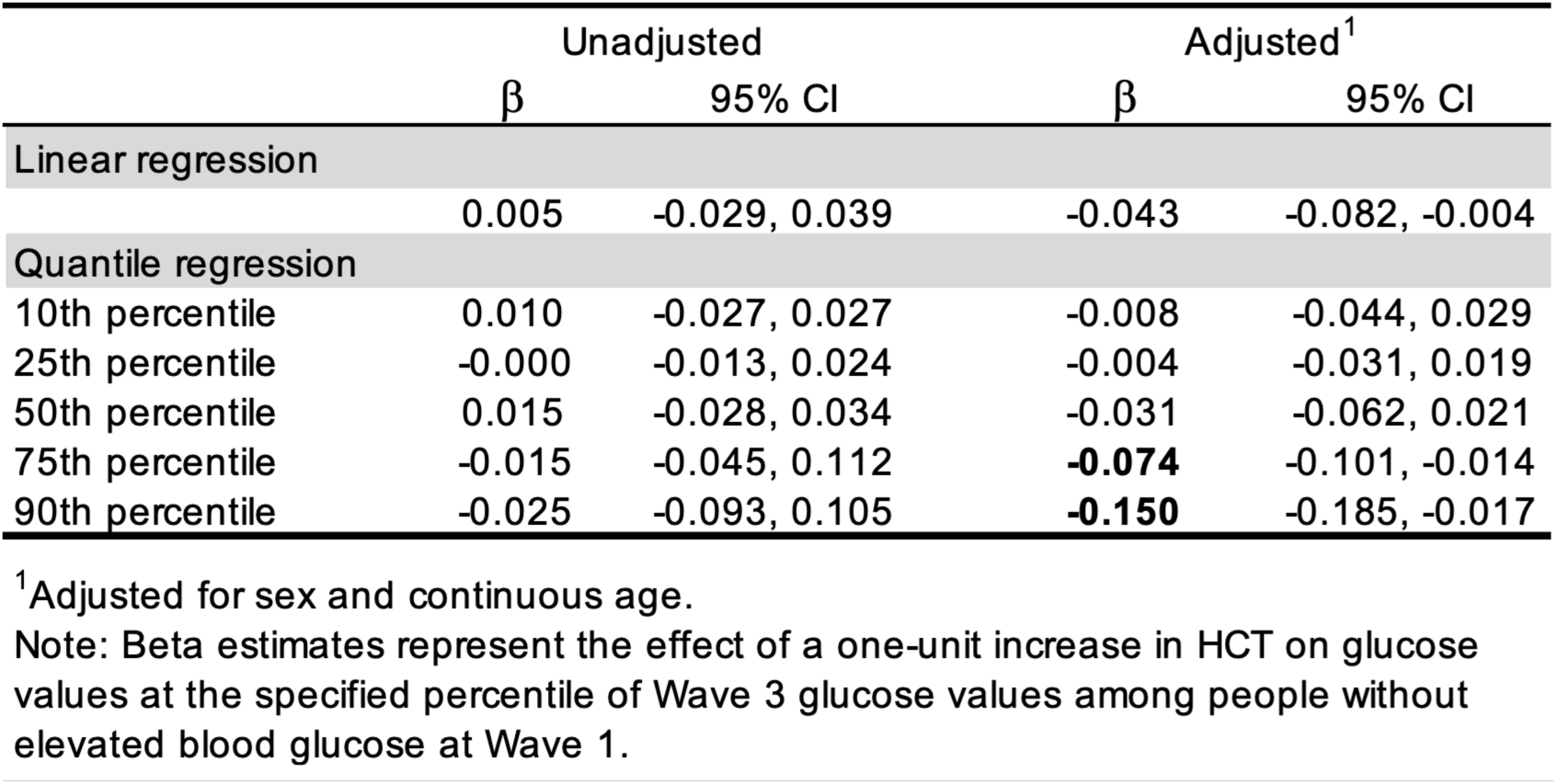
Quantile regression of blood glucose values on HCT.

### Sensitivity analyses

When using WHO criteria to define prediabetes, point estimates and confidence intervals were nearly identical to our primary analysis, indicating that our analysis was robust to choice of prediabetes thresholds. Our analysis was also generally robust to variations in HCT exposure, with similar point estimates and confidence intervals. The one exception was for total cumulative HCT, which estimated a null association within the prediabetes alone subsample in comparison to the inverse association observed in the other 3 operationalizations.

A sensitivity analysis using multiple imputation by fully conditional specification to address missing glucose values at Wave 1 (n = 414) and Wave 3 (n = 1040) yielded similar results to the main analysis. No associations were observed in the population with any glucose impairment or diabetes alone, although adjusted estimates signaled a reduction of risk with marginal significance. In the population with prediabetes alone, the multiply imputed model agreed with reduction of risk, with point estimates within the confidence interval range of the primary analysis but did not retain statistical significance. Sensitivity analysis results are available in the supplemental materials (S3-S5).

## DISCUSSION

Our results suggest that greater household cash transfer access is associated with reduced cumulative incidence of prediabetes. Reduction in prediabetes risk was consistent across age and sex strata. The magnitude of reduced risk was also generally consistent across age and sex strata, with a reduction of risk of 5-6% for each additional R100 of HCT in most subgroups. However, point estimates signal the potential for an even larger reduction for those in mid-life. We did not observe the same association for cumulative incidence of diabetes. Quantile regression revealed the reduction in glucose values associated with HCT was largest at the highest end of the glucose distribution, implying that cash transfer interventions may be most effective for participants with the most severe impairment, even if they do not drop below the diagnostic threshold for diabetes.

Our findings partially align with the limited available literature on the relationship between cash transfers and glucose impairment. A microsimulation study estimated reduced incidence of glucose impairment due to small annual unconditional cash transfer; however, this study was limited incidence of diabetes only without consideration of prediabetes.^59^ Prevalence studies on conditional cash transfer programs found mixed results, with one reporting a decrease in diabetes prevalence and another reporting no difference in diabetes prevalence between those who received the cash transfer and those who did not.^60,61^ Like our quantile regression, preliminary results from an in-progress randomized trial found a statistically significant reduction in a measure of glucose regulation, HbA1c, of 0.61 percentage points due to a monthly USD500 cash transfer among people who already had diabetes, but the transfer was conditional on attending diabetes education and stress coping sessions.^62^ Key differences in conditionality and size of transfer, prevalence instead of incidence, and severity of glycemic dysfunction in study populations may explain departures from our findings.

One explanation for the relationship between HCT and reduced risk of prediabetes but not diabetes among a sample including those with existing prediabetes, is the natural history of the disease. Prediabetes represents an intermediate stage of glucose impairment characterized by insulin resistance and pancreatic cell dysfunction, with substantial annual progression to type 2 diabetes.^6,63^ Because these glucose abnormalities are already established in prediabetes, preventing progression to diabetes may be more challenging than preventing the initial transition from normoglycemia to prediabetes.^64^

A related explanation may be the size of the household cash transfer. Even if more established glycemic dysfunction can be arrested or reversed through levers resourced by additional household income, the amount needed to move the needle may be larger than the current South African cash transfer structure supports. CSG and OPG are intended to offset the costs of living incurred by their beneficiaries rather than fully cover them.^65^ To illustrate, the monthly CSG benefit is equivalent to 75% of the South African food poverty line, or the minimum cost for essential nutrition alone.^65^ This leaves other necessary expenses associated with raising a child—shelter, clothing, school costs, etc.—untouched. Forty-five percent of the South African population depends on cash transfer programs as a major source of income.^66^ Even pooled at the household level, CSG and OPG benefits may be consumed by basic needs of the household before it can be allocated toward expenditures that provide glycemic benefits large enough to prevent progression of established glycemic dysfunction.

At first glance, the risk reduction in prediabetes only may seem contradictory to our observation that HCT is associated with the largest reductions in glucose values among those at the highest end of the glucose distribution. However, this seeming conflict may be explained by an interplay of mathematical and biological phenomena. The range of values from normoglycemia to prediabetes (threshold of 5.6/7.8 mmol/L fasted/nonfasted) and prediabetes to diabetes (threshold of 7.0/11.1 mmol/L fasted/nonfasted) are quite small when compared to the whole spectrum of diabetic hyperglycemia, which can plausibly (although dangerously) reach 33 mmol/L and higher.^67^ Indeed, the maximum glucose value observed in this study was 30.6 mmol/L. The scale difference in these categorical ranges means that the smaller changes observed at the lower end of the glycemic distribution are enough to allow movement across glucose impairment thresholds but similarly sized or even larger changes at the higher end of the spectrum may not achieve the same threshold-crossing effect. The attenuation of effect size at lower quantiles has been observed in other studies of exposure effects on blood glucose distributions.^68,69^

To provide an order-of-magnitude estimate of the potential public health implications, we have calculated hypothetical projections that suggest relatively modest increases in household cash transfers could meaningfully reduce future diabetes burden and associated healthcare expenditures. Using the cumulative incidence of prediabetes and reduction of prediabetes risk associated with each additional R100 of HCT observed in our sample, the HAALSA Indepth cohort would see around 43 fewer cases of prediabetes over seven years, resulting in conservatively 14 fewer ultimate diabetes cases for each additional R100 of HCT.^6^ In the South African context, public sector costs for diabetes treatment and care were estimated at R11,000 per patient annually in 2018, and a person aged 65 with diabetes could expect to live 12 to 16 more years.^70,71^ We project that preventing 14 diabetes cases within the ∼5000-person HAALSA Indepth cohort alone translates to roughly R1.85 million to R2.46 million in avoided diabetes care costs to the public sector.

Importantly, these estimates include only direct medical costs from diabetes cases which did not materialize due to reduction in prediabetes. Economic value may also be generated from delaying or preventing prediabetes itself due to health gains, reduced healthcare utilization and improved health-related quality of life.^72,73^ Additionally, lowering blood glucose levels, even when levels stay in the diabetic range, can reduce risk of serious and costly diabetic complications such as kidney, eye, and cardiovascular disease.^74^ Formal economic modeling using nationally representative data, inclusive of the full continuum of disease and indirect costs, is an important direction for future research evaluating the broader cost-effectiveness of cash transfer programs.

The strengths of this study include use of a large, population-based cohort served by broadly implemented unconditional cash transfer programs. Use of a longitudinal cohort also allowed for incident measures of diabetes and prediabetes, instead of the prevalent measures used in prior work to assess a cash transfer’s impact on diabetes prevention.^60,61^ The exogenous variation in cash transfer eligibility due to program expansions strengthens inference about the impact of cash transfer access on cumulative incidence of diabetes and prediabetes. Another strength is our inclusion of objective biomarker-indicated glucose measures in addition to self-reported diabetes. This triangulation across multiple definitions mitigates risk of diagnostic access bias common in populations with low healthcare utilization and diagnosis of diabetes, as is common in LMICs.^75^ Additionally, we incorporated multiple sensitivity analyses to test the robustness of our findings and the influence of missing data.

Despite this, several limitations of our analysis are acknowledged. First, we use cash transfer *eligibility* as our exposure, as opposed to cash transfer *receipt*. Although this may introduce some measurement error by overestimating the amount of cash transfer income that some households actually receive, it has the benefit of reducing possible confounding due to personal characteristics related to differential uptake, approximating an intent-to-treat analysis and providing policy-relevant estimates of the impact of implementing a cash transfer program.^76^

We use a single reading of blood glucose to assign biomarker-indicated diabetes and prediabetes, but repeated abnormal readings are typically necessary for clinical diagnosis due to normal variability in blood sugar disruptions.^40^ As such, our assignments are more accurately “presumed” statuses. Specific to assignment of prediabetes, there are no universally accepted thresholds for prediabetes using blood glucose measurements. To address the possibility of associations being due to our choice of prediabetes criteria, we performed a sensitivity analysis using different criteria and found our observations robust to this specification. We also performed a quantile regression which leaves blood glucose values as continuous rather than categorized based on pre-defined threshold levels. Nonetheless, future studies should explore using other measures and definitions of glucose impairment such as HbA1c.

Informative censoring due to study drop-out, mortality, or refusal of measurement is possible. For example, rapid developers of diabetes may be more likely to die between study intervals without their outcome being observed. To help mitigate risk of selection bias due to informative censoring, we applied inverse probability of mortality, attrition, and refusal of measurement weights to minimize the likelihood of biased results due to differential missingness.^77^ However, the possibility of selection bias due to pre-baseline mortality or attrition remains.

Finally, we have identified potential limitations in external and internal validity. Due to the unique historical context and characteristics of the Agincourt research area, such as lived experience under South African Apartheid, the results of this study may not be fully generalizable to other LMIC populations. Additionally, we had limited statistical power to detect differences within our subsamples, particularly among the oldest age group. Null associations in subgroups may be due to small sample sizes rather than a true lack of relationship. Future studies should prioritize larger sample sizes to distinguish true null associations from lack of precision.

This study is among the first to examine the relationship between cash transfer programs and cumulative incidence of diabetes and prediabetes in an LMIC population. Overall, evidence suggests that household cash transfer access reduces risk of prediabetes but does not reduce the risk of diabetes. However, household cash transfer access appears to have the largest impact on glucose levels for those at the higher end of the glucose distribution, implying the potential for HCT to partially reduce risk of hyperglycemic complications even if falling short of reaching diabetes thresholds. Future work is needed to identify which lifestyle levers are most responsive to the impact of cash transfer on reducing risk of prediabetes, and if larger or targeted cash transfer amounts could more fully extend this impact to risk of diabetes as well.

## Data Availability

Public use data from Waves 1-3 of the Health and Ageing in Africa: Longitudinal Studies in South Africa (HAALSA) Indepth cohort is available https://www.icpsr.umich.edu/web/NACDA/studies/36633. Agincourt Health and socio-Demographic Surveillance System data can be requested at https://www.agincourt.co.za/data.

https://www.icpsr.umich.edu/web/NACDA/studies/36633

## Funding acknowledgement

This publication is supported by the US National Institute on Aging, NIH (R01AG069128, PIs Rosenberg and Kobayashi) and the US National Center for Advancing Translational Sciences, NIH (TR004389, PIs Hurley and McDowell). The content is solely the responsibility of the authors and does not necessarily represent the official views of the National Institutes of Health.

The Health and Ageing in Africa: Longitudinal Studies in South Africa study is supported by the US National Institute on Aging, NIH (P01AG041710). The HAALSA-HAALSI study is nested within the SAMRC/Wits University Rural Public Health and Health Transitions Research Unit (Agincourt Health and socio-Demographic Surveillance System), supported by the University of the Witwatersrand, Medical Research Council, and Dept of Science and Innovation, South Africa.

## SUPPLEMENTAL MATERIALS

**S1.**
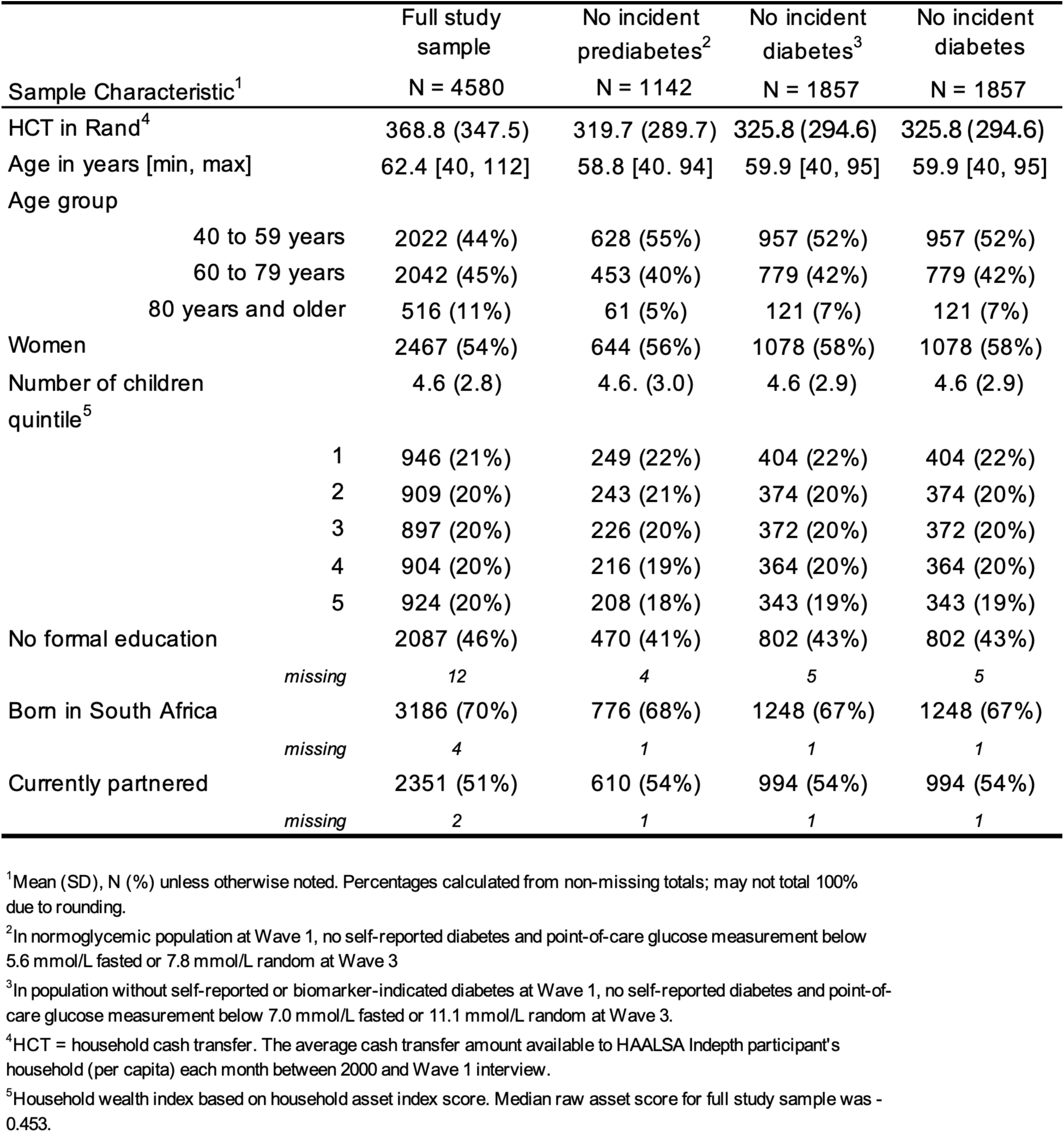
Characteristics of study sample participants who were at risk for but who did not develop prediabetes or diabetes.

**S2.** Probability of no difference in distribution of selected sociodemographic characteristics among participants with and without each incident outcome.

| Sample Characteristic | Incident glucose impairment/<br>No incident glucose impairment | Incident prediabetes/<br>No incident glucose impairment | Incident diabetes/<br>No incident diabetes |
| --- | --- | --- | --- |
| HCT in Rand | 0.966 | 0.906 | 0.424 |
| Age in years [min, max] | **<0.001** | **<0.001** | 0.117 |
| Age group | **<0.001** | **0.013** | 0.132 |
| Women | 0.459 | 0.814 | 0.239 |
| Number of children | 0.085 | 0.414 | **0.041** |
| Household wealth index quintile | 0.308 | 0.257 | 0.069 |
| No formal education | **0.048** | **0.039** | 0.854 |
| Born in South Africa | 0.704 | 0.276 | 0.188 |
| Currently partnered | 0.245 | 0.130 | 0.691 |

Note: P values calculated using Welch's t-test for continuous variables, Mann-Whitney U test for ordered categorical variables, and Pearson chi-square test for unordered categorical variables.

**S3.** Sensitivity analysis: association of HCT with cumulative incidence of glucose impairment using World Health Organization criteria for definition of prediabetes.

|  | ADA thresholds<sup>1</sup> |  |  | WHO thresholds<sup>1</sup> |  |  |
| --- | --- | --- | --- | --- | --- | --- |
|  | CIR | 95% CI | P value | CIR | 95% CI | P value |
| Any glucose impairment |  |  |  |  |  |  |
| Unadjusted | 1.01 | 0.98, 1.03 | 0.659 | 1.01 | 0.98, 1.03 | 0.704 |
| Adjusted<sup>2</sup> | 0.98 | 0.94, 1.02 | 0.282 | 0.97 | 0.93, 1.02 | 0.256 |
| Diabetes alone |  |  |  |  |  |  |
| Unadjusted | 1.01 | 0.96, 1.06 | 0.788 |  |  |  |
| Adjusted<sup>2</sup> | 1.00 | 0.93, 1.07 | 0.908 |  |  |  |
| Prediabetes alone |  |  |  |  |  |  |
| Unadjusted | 0.99 | 0.96, 1.02 | 0.484 | 0.99 | 0.96, 1.03 | 0.647 |
| Adjusted<sup>2</sup> | **0.94** | **0.90, 0.98** | **0.007** | **0.94** | **0.90, 0.99** | **0.014** |

<sup>1</sup>ADA threshold: 5.6 mmol/L. WHO threshold: 6.1 mmol/L
<sup>2</sup>Adjusted for sex and age

**S4.**
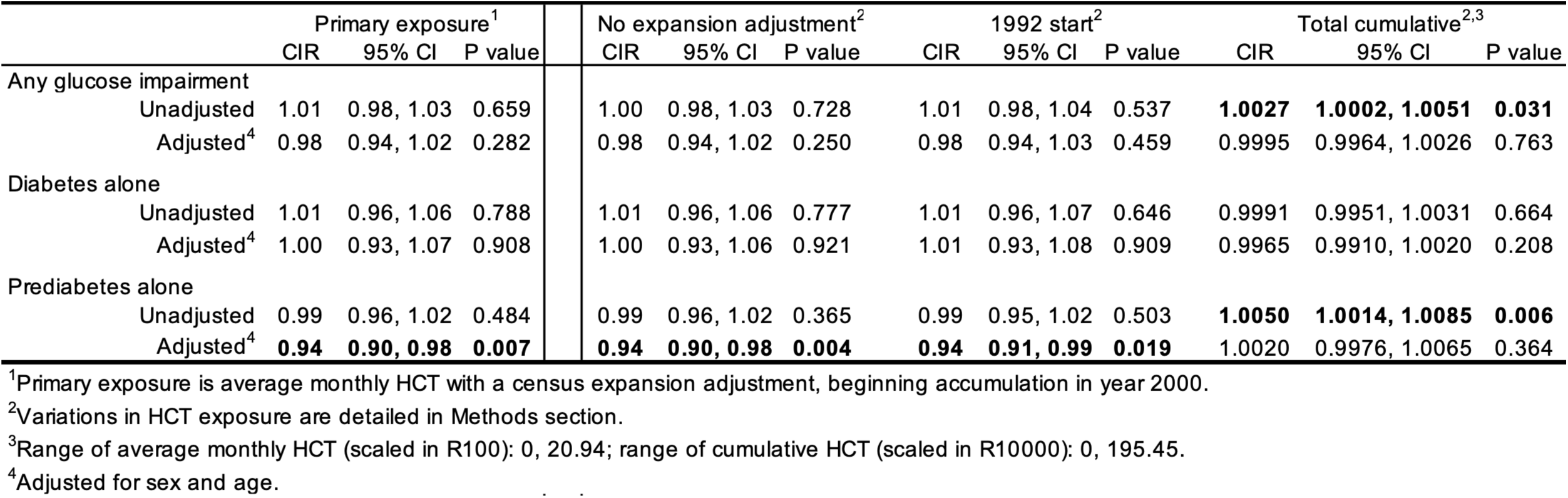
Sensitivity analysis: association of HCT with cumulative incidence of glucose impairment using variations in HCT exposure.

**S5.** Sensitivity analysis using fully conditional specification multiple imputation to address missing glucose values at Wave 1 and Wave 3.

|  | COMPLETE CASE |  |  | MULTIPLY IMPUTED |  |  |
| --- | --- | --- | --- | --- | --- | --- |
|  | CIR | 95% CI | P value | CIR | 95% CI | P value |
| Any glucose impairment |  |  |  |  |  |  |
| Unadjusted | 1.01 | 0.98, 1.03 | 0.659 | 1.01 | 0.98, 1.03 | 0.648 |
| Adjusted<sup>1</sup> | 0.98 | 0.94, 1.02 | 0.282 | 0.98 | 0.95, 1.00 | 0.096 |
| Diabetes alone |  |  |  |  |  |  |
| Unadjusted | 1.01 | 0.96, 1.06 | 0.788 | 0.99 | 0.95, 1.02 | 0.590 |
| Adjusted<sup>1</sup> | 1.00 | 0.93, 1.07 | 0.908 | 0.96 | 0.92, 1.01 | 0.090 |
| Prediabetes alone |  |  |  |  |  |  |
| Unadjusted | 0.99 | 0.96, 1.02 | 0.484 | 1.01 | 0.98, 1.04 | 0.663 |
| Adjusted<sup>1</sup> | **0.94** | **0.90, 0.98** | **0.007** | 0.98 | 0.94, 1.01 | 0.215 |

<sup>1</sup>Adjusted for sex and age

